# Distress and factors for maintaining good mental health among general practitioners during the SARS-CoV-2 pandemic: Results from the cross-sectional PRICOV-19 study in Austria

**DOI:** 10.1101/2024.04.30.24306629

**Authors:** Kathryn Hoffmann, Florian O. Stummer, Esther van Poel, Sara Willems, Silvia Wojczewski

## Abstract

**Background:** The COVID-19 pandemic has had a significant negative impact on the physical and mental health of healthcare workers worldwide. The aim of the paper is to measure the frequency of distress and wellbeing among general practitioners (GPs) in Austria during the pandemic and to identify key levers that could mitigate the risks of distress.

**Methods:** Data were collected as part of the international PRICOV-19 study. In Austria, 500 GPs were randomly selected for participation in a survey between December 2020 and July 2021. For analysis, all dependent and independent variables were described using descriptive statistical methods. Subgroup analyses were conducted using cross-tables and Fisher’s exact tests. A binary logistic regression model was also applied. Open text question was analysed via content analysis.

**Results:** In total, 130 GPs completed the relevant questions for this analysis of the online survey. More than 40% of GPs felt burned out or stated that their work schedules did not leave enough time for personal/family life. Half of the GPs were found to be in distress, with 14.3% in (very) strong distress. More than 40% of the respondents thought that government support was insufficient for the proper functioning of their practice. Working in rural areas was a protective factor against distress, as were sport and exercise, particularly outdoor activity. Connecting with family and friends and adjustments to the work environment to reduce workload were shown to be important.

**Discussion:** Our results show that GPs in Austria suffered from distress during the first two years of the pandemic. To protect GPs as our first-line healthcare workers in pandemic or high-stress situations, several factors are required for a functioning healthcare system: support of GPs regarding work-life balance, support in terms of collaboration between colleagues and the team and easy access to green outdoor spaces for sports and exercise. By identifying key factors that promote good mental health among GPs, healthcare organizations and policymakers can take targeted action to alleviate the negative impact of stress and burnout on this critical sector of the healthcare workforce.

## Background

The COVID-19 pandemic has profoundly affected the physical and mental wellbeing of all demographic groups, with healthcare professionals being particularly susceptible due to their elevated risk of contracting the virus, heightened work-related stress and concerns about transmitting the infection to family members [1-3]. Even before the pandemic, healthcare workers were recognised as a group susceptible to mental health and wellbeing challenges (such as burnout) stemming from factors like high stress levels and long working hours [1]. Symptoms experienced by healthcare workers include depression, depersonalisation, emotional fatigue, diminished sense of personal achievement, anxiety, stress and cognitive and social issues. These symptoms not only directly influence healthcare workers but also their patients, putting the sustainability of the healthcare system at risk. Among physicians reporting at least some burnout symptoms, those in family and emergency medicine are at the greatest risk [4].

The pandemic has heightened the risk of distress for general practitioners/family practitioners (GPs) as frontline workers. Although some researchers advocate addressing both the individual and work environment to mitigate burnout risks and foster wellbeing in the medical field, studies tend to emphasise reducing stress’s negative effects rather than bolstering protective mechanisms, such as emotional and social support [1]. Additionally, the pandemic’s impact on GPs’ wellbeing remains largely unexplored. GPs are known for managing their own physical and mental health in response to the implicit expectations of patients and colleagues to appear healthy and well, as a GP’s health is often seen as a reflection of their medical proficiency [4].

The multi-country analyses of the PRICOV-19 database by Collins et al. [5] described the inter-country variation in the distress levels of GPs across Europe and Israel during the SARS-CoV-2 pandemic. Hereby the Mayo Clinic Wellbeing Index (eMCWI) [6] was used to measure distress scores. The results indicated that the distress levels of Austrian GPs, as represented by box-plot analysis, were comparatively low when compared to the distress levels of GPs in other countries included in the study. For this reason, we decided to take a deeper look at the Austrian situation.

Austria’s healthcare system is grounded in the Bismarkian insurance model, where GPs are small- and medium-sized entrepreneurs and are therefore self-employed. In the public system, they have contracts with social health insurance companies through which services, mainly fee-for-service, are billed directly between GPs and insurance companies. There are now three different forms of organisation: single-handed practices, which account for the largest share (over 80%), group practices and primary care centres and networks. The population in Austria makes frequent use of the outpatient healthcare system, both of GPs and, in almost equal numbers, of specialists working in the outpatient system, since there is no gatekeeping system [7-10]. During the SARS-CoV-2 pandemic, primary care settings such as GPs, a dedicated telephone hotline, and the GP on-call duty service (“Ärztefunkdienst”), were the first points of contact for patients with SARS-CoV-2 infection in Austria. In the first wave in early 2020, patient numbers were slightly lower than normal due to the instructions of public health authorities to stay at home if possible. However, since summer 2020, there has been a sharp increase in the number of patients seen by GPs. GPs must cope with very high demands as in other countries [11, 12]: in addition to the infectious patients, they must safely care for chronically ill patients, continue preventive examinations, see seriously ill patients who have been discharged early from the hospital due to a lack of beds, and remain involved in the testing and vaccination strategies as soon as they become available. In addition, the frequency of telephone consultations increased significantly. Since summer 2020, an increasing number of patients with long-Covid symptoms have been consulting their GPs too [13, 14].

The aim of this study was to assess associations between personal, practice and surrounding factors regarding the distress of Austrian GPs as measured by the eMCWI during the first two years of the SARS-CoV-2 pandemic. We also qualitatively assessed the factors described by the GPs in an open text question in terms of what was good for maintaining their mental health to compare it with the results of the quantitative assessment.

## Methods

### Design

The data were gathered through the PRICOV-19 research project, which was a cross-sectional analysis utilising an online survey distributed to GPs in 37 European nations and Israel. Over 4700 GPs were involved in the research. The PRICOV-19 investigation examined the organisation of GP practices during the SARS-CoV-2 pandemic to ensure secure, efficient, patient-focused and fair treatment. Furthermore, the study explored changes in roles and responsibilities, as well as staff wellbeing [15]. The research was structured following the STROBE guidelines for cross-sectional investigations [16] and was deemed ethically sound by the Medical University of Vienna’s ethics committee (EC N°2200/2020).

### Recruitment

The complex Austrian recruitment strategy was described in detail in a publication by Stummer et al. [17]. We randomly selected, in several steps, 500 GPs from a list provided by the Austrian Chamber of Physicians. The GPs were stratified by sex and county of work to obtain an approximation of the actual national situation. We electronically invited these GPs to participate in this study between December 2020 and July 2021. The invitation contained a brief overview of the research and a hyperlink to the online survey. In total, four reminders were sent. The Research Electronic Data Capture (REDCap) platform [18] was employed to accommodate the questionnaire in all languages, distribute invitations to the national GP practice samples and securely store participants’ responses. Of the 195 Austrian GPs who initiated the online questionnaire, 176 partially completed it (return rate 35.2%). For this particular analysis, 130 responses could be utilised.

### Questionnaire

The PRICOV-19 survey was created in several stages, with a pilot study conducted in Belgium. The final version comprised 53 questions, categorised into six sections: patient flow (appointments, triage and routine care management), infection prevention, information handling, communication, collaboration and self-care, and practice and participant features [15].

National coordinators translated the questionnaire into the languages of the countries involved. In Austria, a group of GPs and colleagues from the Medical University of Vienna (see acknowledgements) assessed the translated survey’s readability and feasibility, after which it was translated back into English. Most important for this analysis were the questions regarding the dependent variables, using the questions of the expanded version of the Mayo Clinic Wellbeing Index (eMCWI) [6].

The eMCWI consists of seven yes/no questions.

During the past month…

1. Have you felt burned out from your work? 2. Have you worried that your work is hardening you emotionally? 3. Have you often been bothered by feeling down, depressed or hopeless? 4. Have you fallen asleep while sitting inactive in a public place? 5. Have you felt that all the things you had to do were piling up so high that you could not overcome them? 6. Have you been bothered by emotional problems (such as feeling anxious, depressed or irritable)? 7. Has your physical health interfered with your ability to do your daily work at home and/or away from home?

Additionally, there were two questions with answers on a Likert scale ranging from 1 (strongly disagree) to 7 (strongly agree).

1. Since the COVID-19 pandemic, the work I do has become more meaningful to me. 2. Since the COVID-19 pandemic, my work leaves me enough time for my personal/ family life.

For these two questions, we clustered the answer options into three groups: (strongly) disagree, neutral and (strongly) agree.

Finally, a distress/wellbeing score was built: cases missing the outcome variable of interest, the eMCWI data, were excluded. “Within the eMCWI, seven items are responded to with a yes (scored as 1) or no (scored as 0), and the remaining two items are responded to on a 7-point or 5-point Likert scale, with those responding strongly disagree/disagree having one point added to their score, those who responded agree/strongly agree having one point subtracted from their score and no adjustment made for those with middle neutral responses. Being at risk of distress is defined as a score of ≥2, as per previous studies” [5].

In addition, the following questions regarding the practice were relevant for the independent, exploratory variables, as they were already in the analysis for the publication by Stummer et al. [17]:

⍰ Work experience: “How many years of work experience do you have in general practice?” The work experience variable was clustered into three categories: 0 years to 4 years and 11 months, 5 years to 14 years and 11 months, and 15 years or more.
⍰ Number of GPs at the practice: “How many GPs and GP trainees are working at this practice? Count every GP and GP trainee as one, irrespective of whether they are full-time or not. Do not forget to include yourself.” The number of GPs working was also grouped into three categories: single-handed (1 GP), 2–3 GPs, and 4 or more GPs.
⍰ Location of the practice: “How would you characterise the location of this practice?” The answer options were big (inner) city, suburbs, (small) town, mixed urban–rural, and rural. The practice location was clustered into big cities, medium-sized locations (i.e., suburbs, small towns, and mixed), and rural areas.
⍰ Size of the patient population: “We would like to get an idea of the size of this practice. How many patients are registered at this practice? If there are no registrations, please indicate the total practice population.” The size of the patient population was clustered into four groups: 0–1999 patients, 2000– 4,999 patients, and 5000 patients or more.

We considered the following surrounding factors particularly relevant for our analysis, since the availability of infection protection equipment, sufficient time for self-education regarding the new medical challenges, the distribution of work in case of co-worker absence, limitations in work due to the infrastructure of the office building, and the absence of governmental support might contribute substantially to the perception of distress for the GPs.

⍰ Infection protection: “Is there enough personal protective equipment (including FFP2/N95 masks) since the pandemic?” The answer options were always, sometimes or never.
⍰ Information processing time: “Is there enough protected time for GPs for reviewing guidelines and going through relevant and reliable literature?” The answers to this question were strongly disagree, disagree, neutral, agree, strongly agree, and don’t know. These options were clustered into the groups (strongly) disagree, neutral, and (strongly) agree; all other answers were counted as missing.
⍰ Distribution of work in the case of co-workers’ absence: “If staff members are absent because of COVID-19 (sick, isolated, quarantined), can the work be distributed in such a way that the wellbeing of colleagues is not compromised?” The answer options were strongly disagree, disagree, neutral, agree, strongly agree, and don’t know. These options were clustered into the groups (strongly) disagree, neutral and (strongly) agree, all other answers we counted as missing.
⍰ Infrastructure concerns: “Since the COVID-19 pandemic, did you experience any limitations related to the building or the infrastructure of this practice to provide high-quality and safe care?” The answers were to a large extent, to a limited extent, hardly, none and don’t know. We dichotomised the answers into “to a large/limited extent” and “hardly/none”, all other answers we counted as missing.
⍰ Governmental support: “Adequate support is provided by the government for the proper functioning of this practice?” The answers to this question were strongly disagree, disagree, neutral, agree, strongly agree, and don’t know. These options were clustered into the groups (strongly) disagree, neutral, and (strongly) agree; all other answers we counted as missing.

### Data analyses

First, descriptive statistical methods (frequencies and percentages) were utilised to characterise all dependent and independent variables. Following this, subgroup analyses were performed using cross-tables and Fisher’s exact tests, which were deemed suitable due to the presence of small sample sizes in several subgroups. A binary logistic regression model was also employed. In this model, the dichotomised eMCWI served as the dependent variable. All explanatory variables were incorporated into the model simultaneously. The outcomes of each regression model are displayed as odds ratios accompanied by 95% confidence intervals. To assess the model fit, Nagelkerke’s R^2^ was provided.

To analyse the open text question regarding how GPs maintain their mental health, we counted the number of participants who provided a text answer to this question, as well as the number of answers per person. The average number of answers was then calculated. The answers were translated into English and analysed with regard to their content and clustered into groups and subgroups. Finally, a word cloud was built with the free word cloud generator available at https://www.wortwolken.com/.

### Ethical considerations

The study was deemed safe by the ethics committee of the Medical University of Vienna (EC N°2200/2020). All participants were classified as experts, and the survey was designed to be completely anonymous. All participants had to read the informed consent at the beginning of the questionnaire and could only continue if they clicked the “I agree” button.

## Results

In total, 130 GPs answered the above-mentioned questions in the 53-question online survey.

### GPs levels of distress during Covid-19

Details of all dependent and independent variables are shown in Table 1. The number of GPs who felt burned out was over 40%, as was the number of GPs who stated that their work schedules did not leave enough time for personal/family life. For the dichotomised eMCWI score, half of the GPs were found to be in distress, and 14.3% were in (very) strong distress (scores 5–9). More than 40% of the respondents did not think that the support from the government was sufficient for the proper functioning of their practice.

**Table 1:**
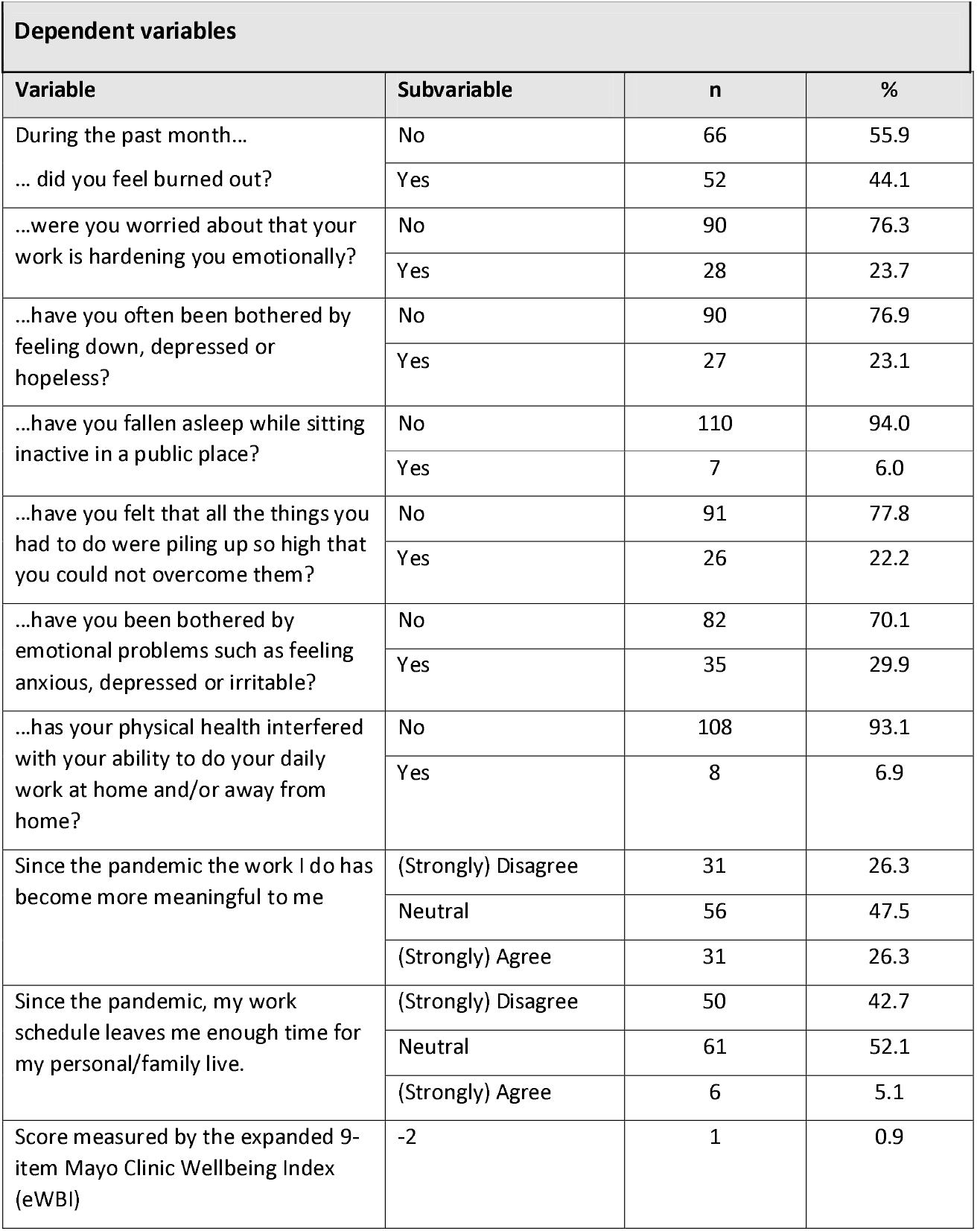
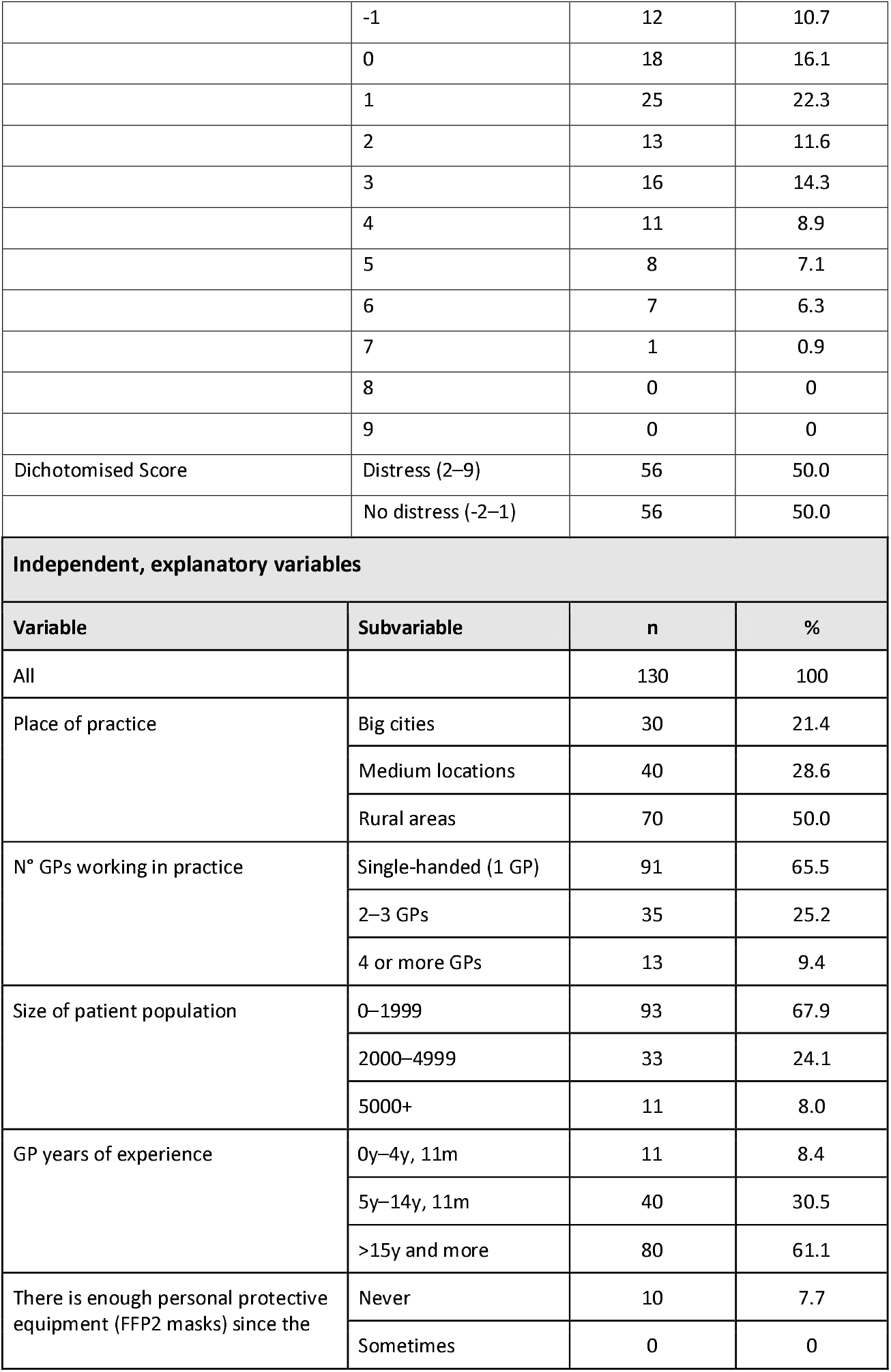
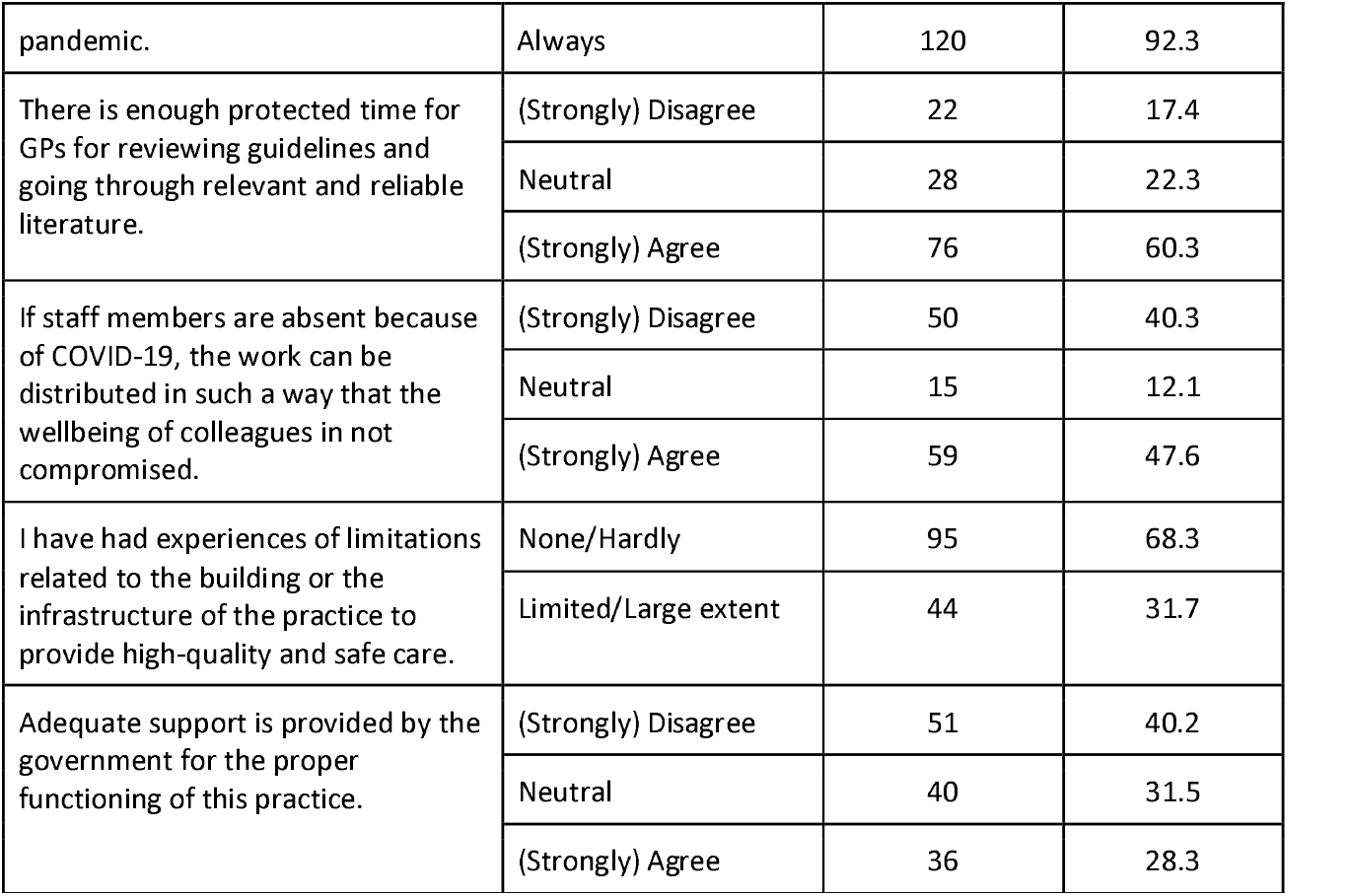
Description of the variables.

Findings of the cross-table calculations in Table 2 show that there is an association between experiencing distress and having between five and 15 years of experience as a GP compared to GPs with 15 years of experience and more. When the distribution of work in the absence of co-workers compromises the wellbeing of colleagues, this is positively associated with distress too.

**Table 2:** Findings of the cross-table calculations.

| Variable | Subvariable | Distress | No distress |
| --- | --- | --- | --- |
|  |  | % (n) | % (n) |
| Place of practice | Big cities | 63.0 (17) | 37.0 (10) |
|  | Medium locations | 46.9 (15) | 53.1 (17) |
|  | Rural areas | 45.3 (24) | 54.7 (29) |
| p |  | 0.326 |  |
| N° GPs working in practice | Single-handed (1 GP) | 52.9 (37) | 47.1 (33) |
|  | 2–3 GPs | 43.3 (13) | 56.7 (17) |
|  | 4 or more GPs | 45.5 (5) | 54.5 (6) |
| p |  | 0.711 |  |
| Size of patient population | 0–1999 | 47.9 (35) | 52.1 (38) |
|  | 2000–4999 | 55.6 (15) | 44.4 (12) |
|  | 5000+ | 50.0 (5) | 50.0 (5) |
| p |  | 0.836 |  |
| GP years of experience | 0y–4y, 11m | 44.4 (4) | 55.6 (5) |
|  | 5y–14y, 11m | 72.2 (26) | 27.8 (10) |
|  | >15y or more | 38.8 (26) | 61.2 (41) |
| p |  | 0.005 |  |
| There is enough personal protective equipment (including FFP2/N95 masks) since the pandemic. | Never | 44.4 (4) | 55.6 (5) |
|  | Always | 50.5 (52) | 49.5 (51) |
| p |  | 0.5 |  |
| There is enough protected time for GPs for reviewing guidelines and going through relevant and reliable literature. | (Strongly) Disagree | 70.0 (14) | 30.0 (6) |
|  | Neutral | 45.8 (11) | 54.2 (13) |
|  | (Strongly) Agree | 47.7 (31) | 52.3 (34) |
| p |  | 0.181 |  |
| If staff members are absent because of COVID-19, the work can be distributed in such a way that the wellbeing of colleagues is not compromised. | (Strongly) Disagree | 64.3 (27) | 35.7 (15) |
|  | Neutral | 27.3 (3) | 72.7 (8) |
|  | (Strongly) Agree | 44.6 (25) | 55.4 (31) |
| p |  | 0.041 |  |
| I have experienced limitations to providing high-quality and safe care related to the building or the infrastructure of the practice. | None/Hardly | 46.1 (35) | 53.9 (41) |
|  | Limited/Large extent | 57.1 (20) | 42.9 (15) |
| p |  | 0.312 |  |
| Adequate support is provided by the government for the proper functioning of this practice. | (Strongly) Disagree | 59.1 (26) | 40.9 (18) |
|  | Neutral | 51.4 (18) | 48.6 (17) |
|  | (Strongly) Agree | 36.4 (12) | 63.6 (21) |
| p |  | 0.144 |  |

When running the binary regression model with all variables included concomitantly (Table 3), two variables remained significantly associated with distress. Compared to big cities, working in a rural area was associated with lower levels of distress (OR 0.21), finally, not having enough time for reviewing guidelines was related to higher distress compared to being neutral.

**Table 3:** Results of the binary regression model.

| Variable | Subvariable | p | OR (95% CI) |
| --- | --- | --- | --- |
| Place of practice | Big cities |  | 1.0 |
|  | Medium locations | 0.05 | 0.27 (0.07–1.02) |
|  | Rural areas | 0.01 | 0.21 (0.06–0.69) |
| N° GPs working in practice | Single-handed (1 GP) |  | 1.0 |
|  | 2–3 GPs | 0.09 | 0.39 (0.13–1.14) |
|  | 4 or more GPs | 0.44 | 0.49 (0.08–3.00) |
| Size of patient population | 0–1999 |  | 1.0 |
|  | 2000–4999 | 0.47 | 1.54 (0.48–4.96) |
|  | 5000+ | 0.62 | 1.57 (0.26–9.42) |
| GP years of experience | 0–4y, 11m |  | 1.0 |
|  | 5y–14y, 11m | 0.27 | 2.86 (0.44–18.75) |
|  | >15y and more | 0.70 | 0.71 (0.13–3.88) |
| There is enough personal protective equipment (including FFP2/N95 masks) since the pandemic. | Never |  | 1.0 |
|  | Always | 0.44 | 2.03 (0.34–12.10) |
| There is enough protected time for GPs for reviewing guidelines and going through relevant and reliable literature. | (Strongly) Disagree |  | 1.0 |
|  | Neutral | 0.04 | 0.19 (0.04–0.92) |
|  | (Strongly) Agree | 0.17 | 0.38 (0.10–1.51) |
| If staff members are absent because of COVID-19, the work can be distributed in such a way that the wellbeing of colleagues is not compromised. | (Strongly) Disagree |  | 1.0 |
|  | Neutral | 0.46 | 0.50 (0.08–3.06) |
|  | (Strongly) Agree | 0.77 | 0.77 (0.25–2.32) |
| I have experienced limitations to providing high-quality and safe care related to the building or the infrastructure of the practice. | None/Hardly | 0.29 | 1.73 (0.63–4.75) |
|  | Limited/Large extent |  | 1.0 |
| Adequate support is provided by the government for the proper functioning of this practice | (Strongly) Disagree |  | 1.0 |
|  | Neutral | 0.78 | 1.17 (0.38–3.63) |
|  | (Strongly) Agree | 0.20 | 0.44 (0.13–1.53) |
| Nagelkerkes R<sup>2</sup> |  |  | 0.324 |

### Factors supporting good mental health

A total of 104 participants answered the open text question about factors that support mental health during the pandemic. Altogether, 231 text answers, mainly words or small sentences, were given. Respondents gave between one and five answers; on average, each person gave two answers.

Table 4 shows the six main categories and related subcategories. Austrian GPs appeared to be quite sporty; only 34 of the 104 participants who answered the question did not list sport or exercise as support in times of crisis. The GPs differentiated clearly between intensive sports and exercise and taking a walk. In the majority of cases, sport and exercise included the addition of “in nature” or “outdoors”. Sometimes, nature was also given as an independent answer. Being outdoors seemed to be important in the inclusion of activities such as gardening, which was stated quite often. Other relevant categories were connecting and talking to special persons, family in particular but also partners, children and/or friends, relaxation, adjusting the work environment and other hobbies like music or reading.

**Table 4:** Six qualitative categories and related subcategories.

| **Sport and exercise** | **Connecting and talking to special persons** | **Nature** | **Relaxation and recovery** | **Adjusting the work environment** | **Other hobbies** |
| --- | --- | --- | --- | --- | --- |
| Sport | Family | Being in nature, outdoors | Meditation | Networking and talking to colleagues | Listening to music |
| Exercise | Children | Sport and exercise in nature | Sleeping | Talking more with the team | Making music |
|  | Partner | Sport and exercise in the fresh air | Active relaxation techniques | CME regarding COVID-19 | Reading (non-medical literature) |
|  | Friends | Gardening | Yoga/Chi-Gong | Supervision | Pets |
|  |  |  | Weekends free | More employees/ substitutes | Others |

## Discussion

Overall, our findings showed that half of the participating GPs in Austria suffered from distress during the first two years of the pandemic. In particular, more than 40% of GPs said that they had feelings of being burned out and not having enough time for personal and/or family life (Table 1). A total of 14.3% were affected by (very) strong distress, meaning every seventh GP (Table 1).

This means that the distress severity score of Austrian GPs might be lower than those of other European countries, according to Collins et al. [5]. However, the frequency of GP distress is far from low at 50%. The international PRICOV-19 publication found that “GPs with less experience, in smaller practices, […] were at a higher risk of distress” [5]. This was not the case for Austria, where neither the number of GPs working in the practice nor the size of the patient population had an association with distress, either in the cross-table results or the binary regression model (Tables 2 and 3). The association between GPs’ experience and distress was inconclusive, since only GPs with working experience between 5 and 14 years and 11 months had significantly less distress than those with less experience. GPs with more than 15 years of experience did not differ significantly from those with little experience. This is partly in contrast with an Italian study, which showed an association between less professional experience and higher levels of anxiety and depression [19]. This could be due to a balancing effect, as Austrian GPs were all equally well equipped (over 94%) with personal protective equipment (PPE), including high-quality masks, regardless of the size of their practice and working experience, and also had fewer problems due to the structural or infrastructural conditions of their practice in an international comparison [20]. The availability of PPI is known to be the most important factor in not only protecting first-line healthcare workers from infectious diseases but also strengthening their mental health, as they are protected from infection themselves and can protect their relatives and friends.

Findings that GPs experienced more distress if it was not possible to distribute work in such a way that the wellbeing of colleagues was not compromised were in line with previous literature (Table 3), although this factor did not maintain significance in the regression model. The probable reasons for distress behind this variable are time pressure issues and an increasing imbalance between work and time for GPs and/or the important people in their lives. This was already experienced by many GPs before the pandemic (Table 1). Another hint for an association between time pressure and distress is in the results of the regression model, in which GPs who did not have enough time for reviewing guidelines and going through relevant and reliable literature had significantly more distress. Time pressure is known to be a stressor that can lead to severe distress and impaired mental health [21-24]. During the SARS-CoV-2 pandemic, time pressure increased, not only because of higher workloads and the need for workplace adjustments, but also due to the need for knowledge about the uncertain infection situation and new treatment options [24, 25].

More than 40% (strongly) disagreed that adequate support was provided by the government for the proper functioning of their practices (Table 1). Governmental support and recognition were found to be important for good mental health in a review of healthcare workers [26] because they contributed to their feelings of being respected and valued [27].

An important finding in our analysis is that what significantly protected Austrian GPs from experiencing distress was working in a rural area (Table 3). This was in contrast to an international study by Petrazzuoli et al. [28], who found that rural practices tend to be smaller and smaller practices tend to be associated with higher distress [29]. However, in Austria, rural practices tend to be larger than those in urban areas because this compensates for lower GP density. Additionally, there are only a few remote areas with a catchment area too small for larger practices.

A plausible explanation for the protecting factor working in rural area, can be found in the analysis of the open text question regarding factors that maintain good mental health for GPs during the pandemic. Most often, GPs indicated that to maintain their mental health, they do sports or exercise, preferably in the fresh air, in nature or simply outdoors (Table 4, Figure 1). It may be that it is easier to go for a walk or run after or between shifts in the countryside. No commuting is necessary; therefore, fewer time constraints occur. Many other studies support the findings that both physical activity and relaxation techniques are helpful for maintaining good mental wellbeing [30].

**Figure 1.**
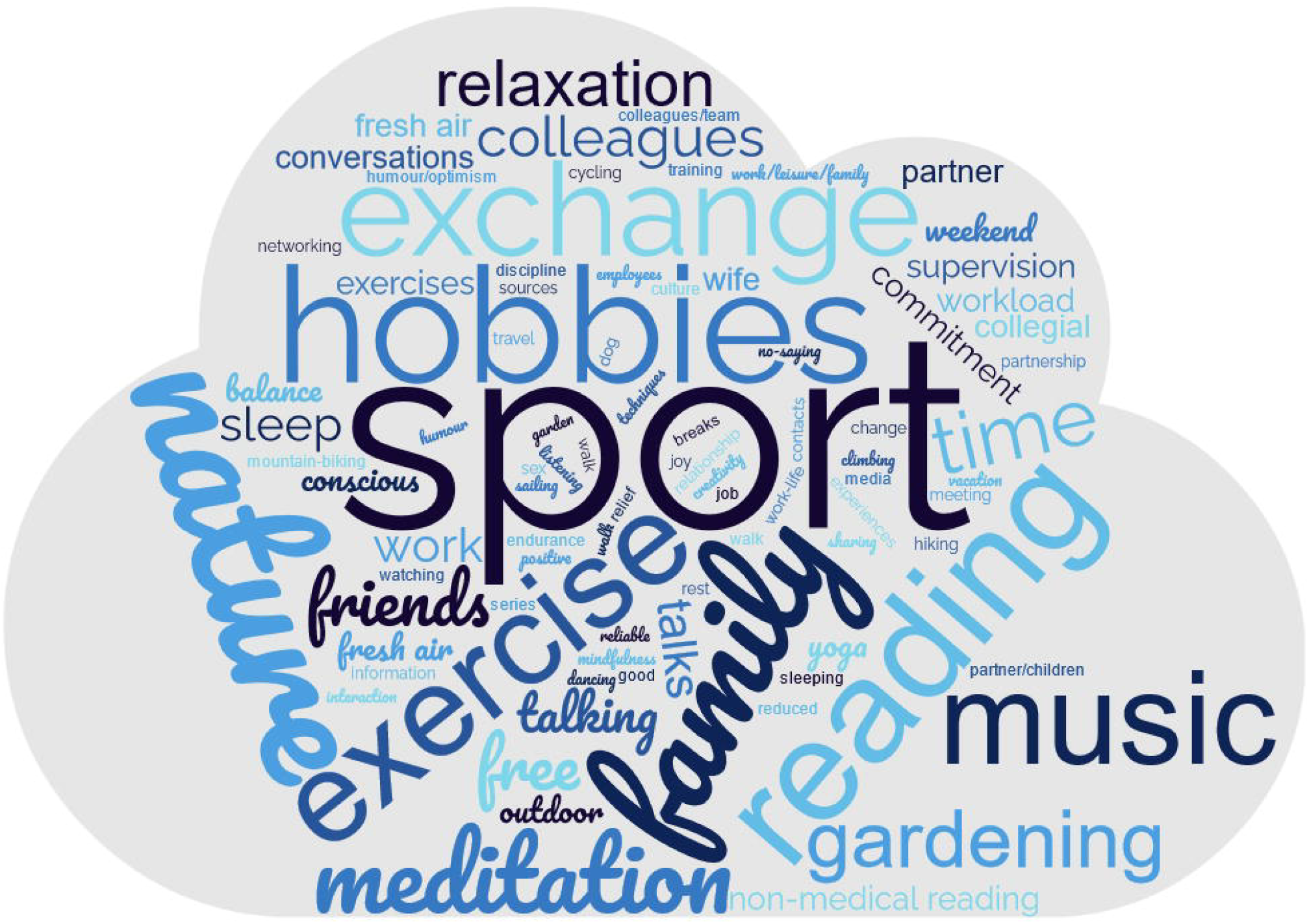
shows the frequency of mentioned words in the word cloud. The ten most frequently mentioned words were sport (n = 54), family (n = 30), exercise (n = 16), hobbies (n = 10), nature (n = 10), reading (n = 9), exchange with friends/colleagues (n = 8), music (n = 8), meditation (n = 6) and gardening (n = 5).

Family and friends, as well as mutual support and regular networking activities, were also noted as important for good mental health. Previous studies have shown that social and emotional support and contact with colleagues and other supporters can reduce mental health issues [31, 32]. Encouraging collaboration between GPs is therefore useful in two ways: it preserves GPs’ mental health and builds up capacity for current and future crises.

### Strengths and limitations

As part of an international project, this study recruited a random sample of GPs in Austria. The development of the questionnaire followed a rigorous protocol: the questionnaire was translated and back-translated, and the questions were culturally adapted. Any discrepancies in terminology were resolved, and the collaborators agreed on a harmonised version with culturally sensitive wording.

The rather low return rate could suggest that a sampling bias exists and that only highly motivated GPs participated, which might have an effect on the external validity of the study or mean that the results are overestimated. On the other hand, it could be that GPs who were heavily involved with COVID-19 had no time to answer the questionnaire and were thus not represented in the study. This, in turn, could have led to an underestimation of the study results. Recall bias is also a common problem in surveys [33]. Given the additional potential for volunteer bias and the cross-sectional survey design [15], direct assessment of causal relationships is not possible.

Another limitation might be that the data collection did not occur directly during the Covid waves. It is not possible to retrospectively estimate the exact Covid burden at a specific point in time. This might have an impact on the accuracy of measuring the impact on wellbeing during the pandemic.

## Conclusion

In conclusion, this study found that approximately half of the participating GPs in Austria experienced distress during the first two years of the pandemic. The most common sources of distress were related to work/free time balance and burnout. However, the severity of distress among Austrian GPs was comparatively lower than in other European countries. Adequate governmental support, fair work distribution and enough specifically dedicated time for reviewing guidelines and the scientific literature were found to be important protective factors against distress.

The strongest protective factor against distress for Austrian GPs was working in a rural area, possibly due to easier access to exercise and outdoor activities in nature. Family, friends, and exchanges and networking with colleagues were also important for maintaining good mental health.

Encouraging collaborative relationships between GPs is necessary to expand their response capacity for current and future crises. Overall, to protect GPs as our first-line healthcare workers in pandemic or high-stress situations, we found several factors that can foster a functioning healthcare system: support of GPs regarding work-life balance, support regarding collaboration between colleagues and the team and easy access to green outdoor spaces for sports and exercise. Free access to hospital or healthcare facility parks and recreational greens for GPs is recommended, especially in urban areas.

## Data Availability

All data produced in the present study are available upon reasonable request to the authors.

## Declarations

### Ethics approval and consent to participate

The study was seen as safe by the ethics committee of the Medical University in Vienna (EC N°2200/2020). All participants were classified as experts, and the survey was designed to be completely anonymous. All participants had to read the informed consent before the start of the questionnaire and proceeded only if they clicked the “I agree” button.

### Consent to publish

The authors confirm (1) that the work described has not been published before, (2) that it is not under consideration for publication elsewhere, (3) that its publication has been approved by all co-authors, and (4) that its publication has been approved (tacitly or explicitly) by the responsible authorities at the institution where the work is carried out.

### Availability of data and materials

All data are centrally stored on the server of Ghent University (Belgium). All data were anonymised at Ghent University, and all raw data that could lead to the identification of the respondents was permanently removed. A reasonable request is required to access non-identifiable data by users who are external to the PRICOV-19 consortium. Access will be subject to a data transfer agreement and following approval from the principal investigator of the PRICOV-19 study.

### Author contributions

KH coordinated the Austrian part of the PRICOV-19 study, analysed the Austrian data and drafted the first version of the manuscript together with FOS and SW. EVP supported the conceptualisation and design of the international study and performed the data cleaning. SW is the international PI of the Pricov19 study. All authors critically reviewed and provided comments on the paper and approved the final manuscript.

## Acknowledgements

We would like to thank Ruth Kutalek, Franz Mayrhofer, Susanne Rabady and Maria Wendler for their support regarding the translation and test of the Austrian questionnaire for the PRICOV-19 study.

## Author information

K Hoffmann has a full professorship in primary care medicine and chairs the Department of Primary Care Medicine at the Medical University of Vienna, Austria. Additionally, she is an MD, master of public health and general practitioner.

FO Stummer is a DBA holder in strategic management and a master of public health, finishing his PhD at the Medical University of Vienna, Austria.

Evan Poel is an MD and PhD student in the Department of Public Health and Primary Health Care at the University of Gent, Belgium.

S. Willems has a full professorship in equity in healthcare, is MD and chairs the Department of Public Health and Primary Health Care at the University of Gent, Belgium

S Wojczewski is a post-doc researcher at the Centre for Public Health at the Medical University of Vienna. She holds a PhD in anthropology from the University of Lausanne, Switzerland.

## References

1. Expert Panel on effective ways of investing in health (EXPH): Opinion on supporting mental health of health workforce and other essential workers. Health and Food Safety Directorate-General. Publications Office of the European Union: Luxembourg; 2021.

2. Saragih ID, Tonapa SI, Saragih IS, Advani S, Batubara SO, Suarilah I, Lin CJ: Global prevalence of mental health problems among healthcare workers during the Covid-19 pandemic: A systematic review and meta-analysis. Int J Nurs Stud 2021, 121:104002.

3. Vanhaecht K, Seys D, Bruyneel L, Cox B, Kaesemans G, Cloet M, Van Den Broeck K, Cools O, De Witte A, Lowet K et al: COVID-19 is having a destructive impact on health-care workers’ mental well-being. Int J Qual Health Care 2021, 33(1).

4. Murray M, Murray L, Donnelly M: Systematic review of interventions to improve the psychological well-being of general practitioners. BMC Fam Pract 2016, 17:36.

5. Collins C, Clays E, Van Poel E, Cholewa J, Tripkovic K, Nessler K, de Rouffignac S, Santric Milicevic M, Bukumiric Z, Adler L et al: Distress and wellbeing among general practitioners in 33 countries during COVID-19: Results from the cross-sectional PRICOV-19 study to inform health system interventions. Int J Environ Res Public Health 2022, 19(9).

6. Dyrbye LN, Satele D, Shanafelt T: Ability of a 9-item well-being index to identify distress and stratify quality of life in US workers. J Occup Environ Med 2016, 58(8):810–817.

7. Bachner F, Bobek J, Habimana K, Ladurner J, Lepuschütz L, Ostermann H: Das österreichische Gesundheitssystem. Akteure, Daten, Analysen. WHO Regional Office for Europe; 2019.

8. Hoffmann K, George A, Jirovsky E, Dorner TE: Re-examining access points to the different levels of health care: a cross-sectional series in Austria. Eur J Public Health 2019, 29(6):1005–1010.

9. Hoffmann K, George A, Van Loenen T, De Maeseneer J, Maier M: The influence of general practitioners on access points to health care in a system without gatekeeping: a cross-sectional study in the context of the QUALICOPC project in Austria. Croat Med J 2019, 60(4):316–324.

10. Hoffmann K, Ristl R, George A, Maier M, Pichlhofer O: The ecology of medical care: access points to the health care system in Austria and other developed countries. Scand J Prim Health Care 2019, 37(4):409–417.

11. Ares-Blanco S, Guisado-Clavero M, Ramos Del Rio L, Gefaell Larrondo I, Fitzgerald L, Adler L, Assenova R, Bakola M, Bayen S, Brutskaya-Stempkovskaya E et al: Clinical pathway of COVID-19 patients in primary health care in 30 European countries: Eurodata study. Eur J Gen Pract 2023:2182879.

12. Rawaf S, Allen LN, Stigler FL, Kringos D, Quezada Yamamoto H, van Weel C, Global Forum on Universal Health C, Primary Health C: Lessons on the COVID-19 pandemic, for and by primary care professionals worldwide. Eur J Gen Pract 2020, 26(1):129–133.

13. Expert Panel on effective ways of investing in health (EXPH): Facing the impact of post-Covid-19 condition (Long COVID) on health systems. Health and Food Safety Directorate-General. Publications Office of the European Union: Luxembourg; 2022.

14. Davis HE, McCorkell L, Vogel JM, Topol EJ: Long COVID: Major findings, mechanisms and recommendations. Nat Rev Microbiol 2023, 21(3):133–146.

15. Van Poel E, Vanden Bussche P, Klemenc-Ketis Z, Willems S: How did general practices organize care during the COVID-19 pandemic: the protocol of the cross-sectional PRICOV-19 study in 38 countries. BMC Prim Care 2022, 23(1):11.

16. von Elm E, Altman DG, Egger M, Pocock SJ, Gotzsche PC, Vandenbroucke JP, Initiative S: The Strengthening the Reporting of Observational Studies in Epidemiology (STROBE) statement: guidelines for reporting observational studies. Ann Intern Med 2007, 147(8):573–577.

17. Stummer FO, Voggenberger L, Gomez Pellin M, Van Poel E, Willems S, Hoffmann K: Insights into the use of telemedicine in primary care in times of the SARS-CoV-2 pandemic. A cross-sectional analysis based on the international PRICOV-19 study in Austria. BMC Fam Pract; Supplement 2023 [in press].

18. Harris PA, Taylor R, Minor BL, Elliott V, Fernandez M, O’Neal L, McLeod L, Delacqua G, Delacqua F, Kirby J et al: The REDCap consortium: Building an international community of software platform partners. J Biomed Inform 2019, 95:103208.

19. Lasalvia A, Rigon G, Rugiu C, Negri C, Del Zotti F, Amaddeo F, Bonetto C: The psychological impact of COVID-19 among primary care physicians in the province of Verona, Italy: a cross-sectional study during the first pandemic wave. Fam Pract 2022, 39(1):65–73.

20. Windak A, Nessler K, Van Poel E, Collins C, Wojtowicz E, Murauskiene L, Hoffmann K, Willems S: Responding to COVID-19: The Suitability of Primary Care Infrastructure in 33 Countries. Int J Environ Res Public Health 2022, 19(24).

21. Matheson C, Robertson HD, Elliott AM, Iversen L, Murchie P: Resilience of primary healthcare professionals working in challenging environments: a focus group study. Br J Gen Pract 2016, 66(648):e507–515.

22. Robertson HD, Elliott AM, Burton C, Iversen L, Murchie P, Porteous T, Matheson C: Resilience of primary healthcare professionals: a systematic review. Br J Gen Pract 2016, 66(647):e423–433.

23. Tsiga E, Panagopoulou E, Sevdalis N, Montgomery A, Benos A: The influence of time pressure on adherence to guidelines in primary care: an experimental study. BMJ Open 2013, 3(4).

24. Galbraith N, Boyda D, McFeeters D, Hassan T: The mental health of doctors during the COVID-19 pandemic. BJPsych Bull 2021, 45(2):93–97.

25. Lingum NR, Sokoloff LG, Meyer RM, Gingrich S, Sodums DJ, Santiago AT, Feldman S, Guy S, Moser A, Shaikh S et al: Building long-term care staff capacity during COVID-19 through just-in-time learning: Evaluation of a modified ECHO model. J Am Med Dir Assoc 2021, 22(2):238–244 e231.

26. De Kock JH, Latham HA, Leslie SJ, Grindle M, Munoz SA, Ellis L, Polson R, O’Malley CM: A rapid review of the impact of COVID-19 on the mental health of healthcare workers: Implications for supporting psychological well-being. BMC Public Health 2021, 21(1):104.

27. Merino MD, Privado J: Does employee recognition affect positive psychological functioning and well-being? Span J Psychol 2015, 18:E64.

28. Petrazzuoli F, Collins C, Van Poel E, Tatsioni A, Streit S, Bojaj G, Asenova R, Hoffmann K, Gabrani J, Klemenc-Ketis Z et al: Differences between rural and urban practices in the response to the COVID-19 pandemic: Outcomes from the PRICOV-19 study in 38 countries. Int J Environ Res Public Health 2023, 20(4).

29. Werdecker L, Esch T: Burnout, satisfaction and happiness among German general practitioners (GPs): A cross-sectional survey on health resources and stressors. PLoS One 2021, 16(6):e0253447.

30. Janssen M, Heerkens Y, Kuijer W, van der Heijden B, Engels J: Effects of Mindfulness-Based Stress Reduction on employees’ mental health: A systematic review. PLoS One 2018, 13(1):e0191332.

31. Muller AE, Hafstad EV, Himmels JPW, Smedslund G, Flottorp S, Stensland SO, Stroobants S, Van de Velde S, Vist GE: The mental health impact of the covid-19 pandemic on healthcare workers, and interventions to help them: A rapid systematic review. Psychiatry Res 2020, 293:113441.

32. Verhoeven V, Tsakitzidis G, Philips H, Van Royen P: Impact of the COVID-19 pandemic on the core functions of primary care: Will the cure be worse than the disease? A qualitative interview study in Flemish GPs. BMJ Open 2020, 10(6):e039674.

33. Peersman W, Pasteels I, Cambier D, De Maeseneer J, Willems S: Validity of self-reported utilization of physician services: A population study. Eur J Public Health 2014, 24(1):91–97.

